# Long term impact on lung function of patients with moderate and severe COVID-19. A prospective cohort study

**DOI:** 10.1101/2021.01.06.21249312

**Authors:** Sonia Qureshi, Nosheen Nasir, Naveed Haroon Rashid, Naveed Ahmed, Zoya Haq, Farah Naz Qamar

**Affiliations:** Department of Paediatrics and Child Health, Aga Khan University, Stadium Road, P.O. Box 3500, Karachi 74800, Pakistan; Department of Medicine, Aga Khan University, Stadium Road, P.O. Box 3500, Karachi 74800, Pakistan; Liaquat National Hospital and Medical College, Karachi, Pakistan

**Author notes:** ***Corresponding author***: Farah Naz Qamar, Associate Professor, Dept. of Paediatrics & Child Health, Aga Khan University Hospital Karachi, Pakistan, Stadium Road P.O. Box 3500, Karachi 74800, Pakistan. Authors Email ID,.

## Abstract

**Introduction:** A significant number of patients continue to recover from COVID-19; however, little is known about the lung function capacity among survivors. We aim to determine the long-term impact on lung function capacity in patients who have survived moderate or severe COVID-19 disease in a resource-poor setting.

**Methods and analysis:** This prospective cohort study will include patients aged 15 years and above and have reverse transcriptase-polymerase chain reaction (RT-PCR) positive for COVID 19 (nasopharyngeal or oropharyngeal). Patients with a pre-existing diagnosis of obstructive or interstitial lung disease, lung fibrosis and cancers, connective tissue disorders, autoimmune conditions affecting the lungs, underlying heart disease, history of syncope and refuse to participate will be excluded. Pulmonary function will be assessed using spirometry and diffusion lung capacity for carbon monoxide (DLCO) at three- and six-months interval. A chest X-ray at three and six-month follow-up and CT-chest will be performed if clinically indicated after consultation with the study pulmonologist or Infectious Disease (ID) physician. Echocardiogram (ECHO) to look for pulmonary hypertension at the three months visit and repeated at six months if any abnormality is identified initially. Data analysis will be performed using standard statistical software.

**Ethics and dissemination:** The proposal was reviewed and approved by ethics review committee (ERC) of the institution (ERC reference number 2020-4735-11311). Informed consent will be obtained from each study participant. The results will be disseminated among study participants, institutional, provincial and national level through seminars and presentations. Moreover, the scientific findings will be published in high-impact peer-reviewed medical journals.

**Strengths and Limitations of this study:**

- The study has the potential to develop context-specific evidence on the long-term impact on lung function among COVID-19 survivors
- Findings will play key role in understanding the impact of the disease on vital functions and help devise rehabilitative strategies to best overcome the effects of disease
- This is a single-center, study recruiting only a limited number of COVID-19 survivors
- The study participants may loss-to-follow up due to uncertain conditions and disease reemergence

## Background

The severe acute respiratory syndrome coronavirus 2 (SARS-CoV-2) also known as COVID-19, was declared as a global pandemic by World Health Organization (WHO) during first quarter of 2020. The virus was traced to the Chinese province of Wuhan where it first emerged in December 2019 (1). Globally, as of November 7, 2020, there are 49,685,311 confirmed cases of COVID-19, including 1,249,030 deaths, reported to WHO (2). The case fatality rate of COVID-19 is lower (2%) in contrast, Severe Acute Respiratory Syndrome (SARS) in 2003 (∼10% CFR) and Middle East respiratory syndrome coronavirus (MERS-CoV) in 2012 (34% CFR) (3, 4). In Pakistan, after implementation of a six-month, nation-wide lockdown the number of cases declines, with daily active cases being 666, on 12^th^ October 2020, as compared to the peak of 6,825, during June 2020 (5). However, a second wave of COVID-19 cases is now being observed throughout the country which is believed to be due to non-compliance of standard operating procedures (SOPs) imposed by the government (6).

Since COVID-19 has a predilection for the lung, concerns have been raised about the restoration of lung function after apparent recovery from the disease. COVID-19 presents with a variable degree of clinical severity, ranging from mild upper respiratory tract illness to severe interstitial pneumonia and acute respiratory distress syndrome (ARDS) (7-9). MERS, Intensive Care Unit (ICU) survivors have been reported to have limitations in some measures which altered their quality of life and led to long term psychological morbidities when compared to patients with less severe illness managed in medical wards (10). The pathophysiological features of severe Covid-19 are dominated by an acute pneumonic process with an extensive radiologic opacity and, on autopsy, diffuse alveolar damage, inflammation, microvascular thrombosis along with capillary congestion in lungs (10). Recovered and discharged patients with COVID-19 pneumonia were seen to have residual abnormalities in chest computed tomography (CT) scans, with ground-glass opacity as the most common pattern (11).

Older patients with co-morbidities such as hypertension, diabetes, malignancies and risk factors such as smoking were found to be at greater risk of severe disease. Comorbidities have been correlated with poorer clinical outcomes (12). In addition to the direct effects of the disease, increased medical cost and decreased capacity to generate income due to severe, prolonged illness might be cumbersome for families. Considering the novelty and unpredictability of the disease, it is critical to study the long-term impact on lung function of patients recovered from the disease.

## Aim

To monitor the lung function of patients with moderate and severe COVID-19 disease at three and six months by performing pulmonary function test at a tertiary care hospital of a low-middle income country.

### Research question

What is the long-term impact of moderate or severe COVID-19 illness on pulmonary functions?

### Research objective

To determine the long-term impact on lung function in patients suffering from moderate and severe COVID-19 disease in a low-middle income country.

## Methods

### Study design and setting

This prospective cohort study will be undertaken at Aga Khan University Hospital (AKUH) Karachi, Pakistan. AKUH is one of the tertiary care hospitals in Karachi Pakistan with inpatient bed capacity more than 700 and a 24/7 operational emergency room. It consists of undergraduate medical and nursing school, and postgraduate training programs. In terms of services, the institute has advanced capacity in almost every specialty such as internal medicine, infectious disease, hematology, and pathology and laboratory services. It is one of the four institutes across Pakistan to initiate COVID-19 testing early in the outbreak and reached to testing capacity of almost > 1000 tests per day. AKUH led the training and guideline development for management of COVID-19 in Pakistan.

### Study population

All patients aged 15 years and above with a PCR confirmed diagnosis of COVID-19, having moderate or severe COVID-19 disease as per WHO criteria with a pre-defined inclusion and exclusion criteria will be enrolled in the study (13).

### Inclusion criteria

1. Patients of age ≥15 years who suffered from moderate or severe illness related to COVID-19 during an outbreak (classified using WHO criteria).
2. RT-PCR positive for COVID 19 (Nasopharyngeal or oropharyngeal).
3. Consent to participate.

### Exclusion criteria

1. Patients suffering from underlying obstructive disease such as asthma, chronic obstructive pulmonary disease (COPD), bronchiectasis or interstitial lung disease, lung fibrosis, lung cancers, connective tissue disorders like scleroderma, autoimmune conditions affecting the lungs like sarcoidosis and Wegener’s disease.
2. Patients with underlying heart disease.
3. Patients with history of syncope.

### Sampling technique

Patients suffering from moderate or severe COVID-19 with positive nasopharyngeal or oropharyngeal RT-PCR will be recruited consecutively from the study site. COVID-19 patients admitted in the ICU or high dependency unit (HDU) will be screened for inclusion in the study. Eligible patients will be approached for inclusion in the study after discharge from the hospital. Participants will be contacted over phone within one month of discharge from hospital and verbal consent will be obtained

Once the consent has been obtained, an appointment for pulmonary function test (spirometry/ DLCO), chest x-ray and echocardiogram (ECHO) will be scheduled at third month post discharge (first follow up). In order to facilitate the patients and avoid overflow in clinical areas, the appointment will be kept flexible with one-week (before or after) window according to availability of patient and appointment slot.

Following the investigations, the patients will be asked to visit for consultation with a pulmonologist or infectious disease consultant to discuss the reports and further advise on the disease. Patients will be thoroughly examined by the consultant and advised further radiology examination if needed. A second follow up will be scheduled at six months for a repeat, pulmonary function test and chest X-ray. ECHO will be repeated only if an abnormality is reported at the 3 months follow up. A six-month follow-up visit with consultant will be scheduled to predict the outcome of the disease (Figure 1). CT chest will be performed at three-month follow-up in patients whose chest X-ray shows persistent abnormality when compared to the baseline shows a new abnormality or on the physicians’ recommendation.

**Figure 1.**
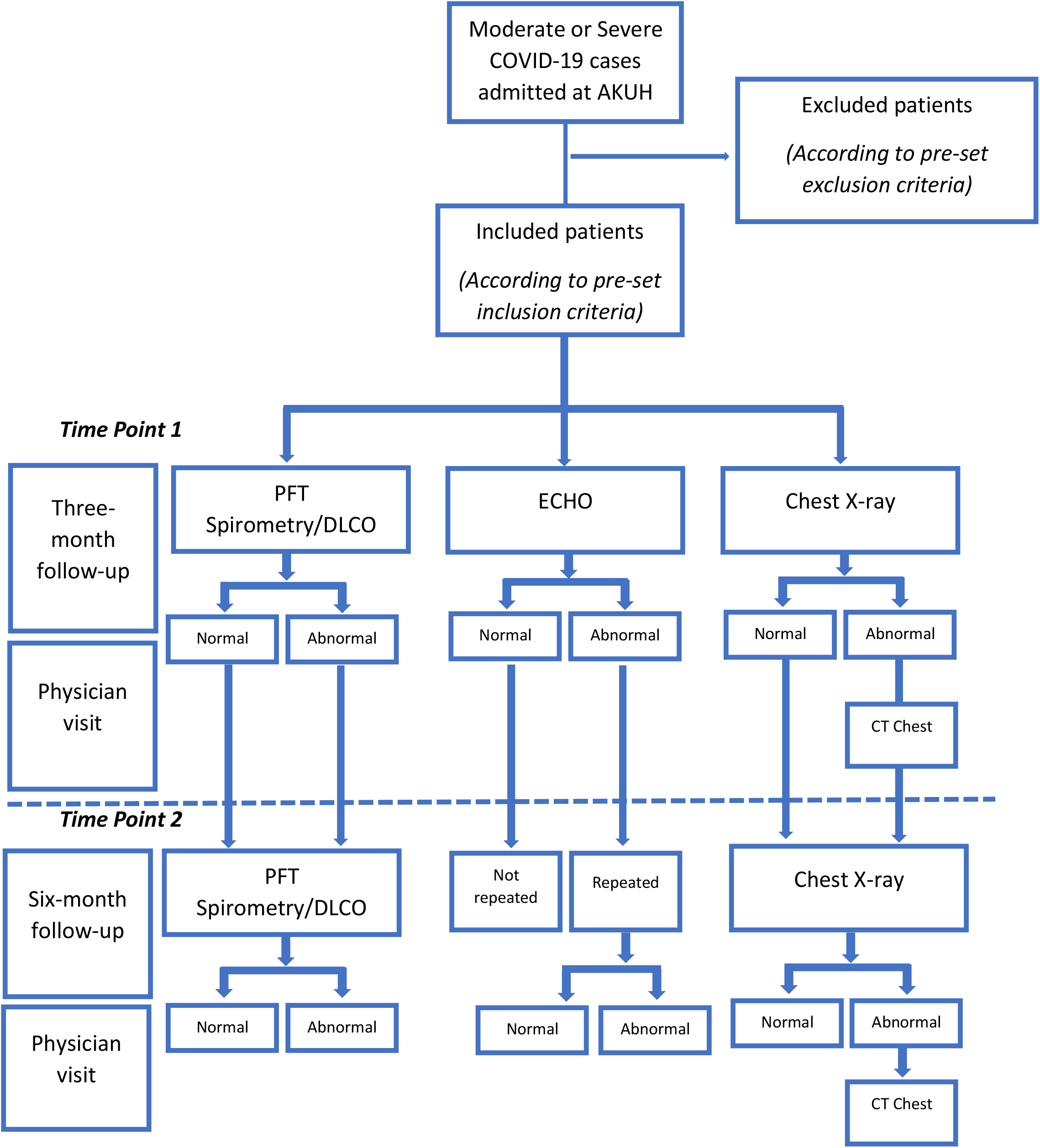
Study flow chart: time points of follow-ups, testing and physician visits.

### Sample size

This will be an exploratory study therefore; we will enroll 60-70 confirmed COVID-19 cases fulfilling the inclusion criteria. Most (80%) of COVID-19 cases are mild, and the number of COVID-19 cases is expected to decline in the next 3 months thus, an arbitrary sample size of 60-70 is selected for convenience and feasibility. This sample may be underpowered to detect the outcome however; it will be helpful in generating the proof of principle.

### Data collection tool

All clinical, epidemiological, demographic and laboratory data will be collected on a predesigned paper-based questionnaire. The questionnaire consists of sections for demographics, clinical and laboratory data. The physician notes will be extracted from the medical record file of the patients. Following study tools will be used to collect data;

1. Screening form for evaluation of eligibility of participants.
2. Questionnaire to record demographics, date of onset of disease, medical history including comorbidities, smoking status, influenza vaccine history, travel history. The information on this form will be collected from several sources including patients, parents/caregivers and hospital records.
3. Laboratory form will be used to record findings of lung function test, Chest X-ray, CT-chest and ECHO findings at 3- and 6-months follow-ups.

### Data collection procedure

The data will be collected from different sources and time points from diagnosis to final follow-up of the study as depicted in (Table 2). Lung function impairment will be measured by performing pulmonary function test (spirometry and DLCO) and reported as a categorical variable, as defined by the guidelines of the American Thoracic Society (ATS). COVID-19 disease will be categorized as moderate or severe/ critical illness based on World Health Organization (WHO) Criteria depicted in (Table 1).

**Table 1:**
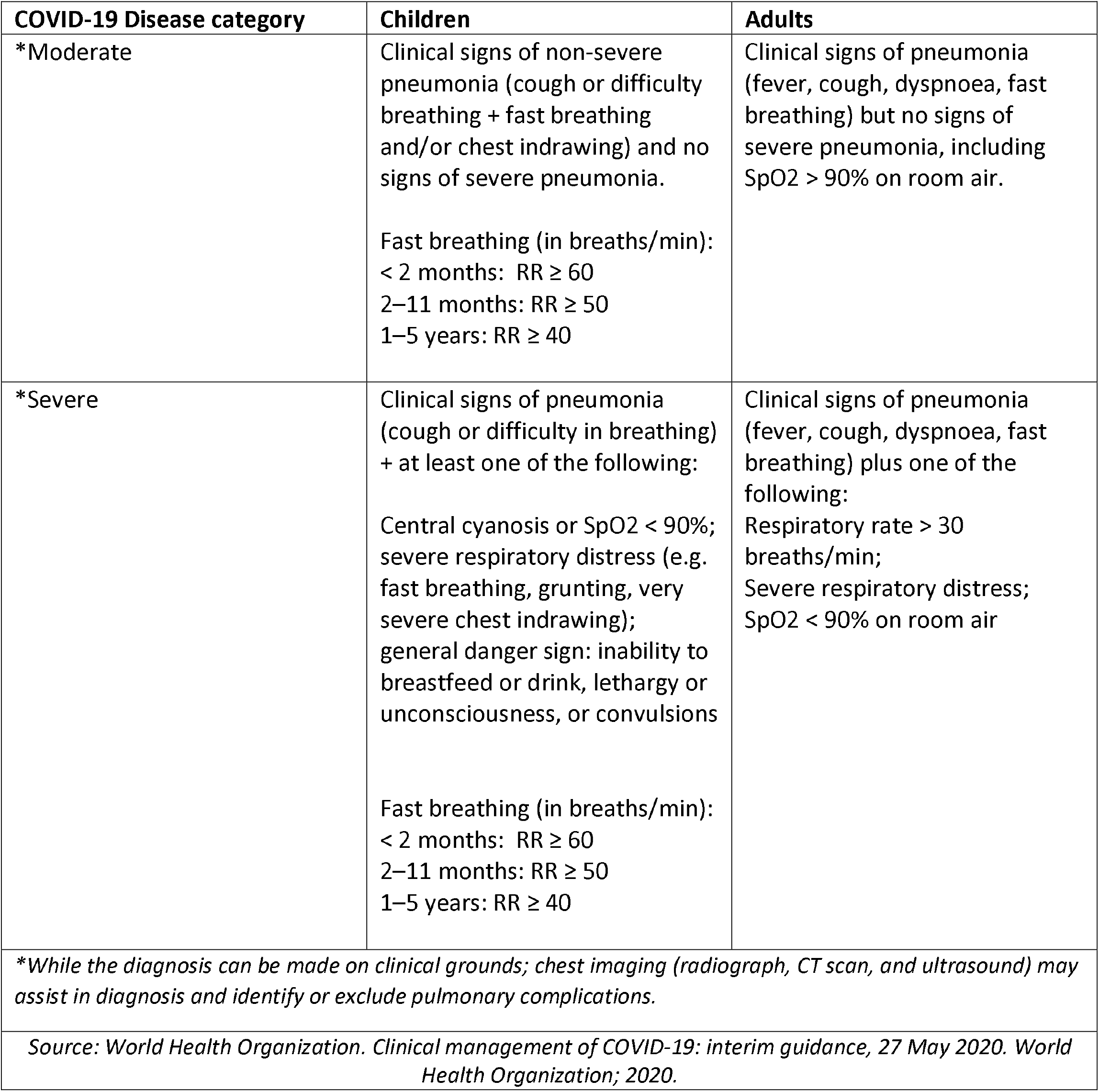
World Health Organization (WHO) -COVID-19 Disease classification.

| COVID-19 Disease category | Children | Adults |
| --- | --- | --- |
| *Moderate | Clinical signs of non-severe pneumonia (cough or difficulty breathing + fast breathing and/or chest indrawing) and no signs of severe pneumonia.<br>Fast breathing (in breaths/min):<br>< 2 months: RR ≥ 60<br>2–11 months: RR ≥ 50<br>1–5 years: RR ≥ 40 | Clinical signs of pneumonia (fever, cough, dyspnoea, fast breathing) but no signs of severe pneumonia, including SpO<sub>2</sub> > 90% on room air. |
| *Severe | Clinical signs of pneumonia (cough or difficulty in breathing) + at least one of the following:<br>Central cyanosis or SpO<sub>2</sub> < 90%; severe respiratory distress (e.g. fast breathing, grunting, very severe chest indrawing); general danger sign: inability to breastfeed or drink, lethargy or unconsciousness, or convulsions<br>Fast breathing (in breaths/min):<br>< 2 months: RR ≥ 60<br>2–11 months: RR ≥ 50<br>1–5 years: RR ≥ 40 | Clinical signs of pneumonia (fever, cough, dyspnoea, fast breathing) plus one of the following:<br>Respiratory rate > 30 breaths/min;<br>Severe respiratory distress;<br>SpO<sub>2</sub> < 90% on room air |
| *\*While the diagnosis can be made on clinical grounds; chest imaging (radiograph, CT scan, and ultrasound) may assist in diagnosis and identify or exclude pulmonary complications.* |  |  |
| *Source: World Health Organization. Clinical management of COVID-19: interim guidance, 27 May 2020. World Health Organization; 2020.* |  |  |

**Table 2:** Study variables and time points for data collection.

| S.NO | Variables | Hospital admission records |  |  |  | Prospective from patient records |  |
| --- | --- | --- | --- | --- | --- | --- | --- |
|  |  | Baseline | 24-48 hours Pre administration of anti-COVID-19 medication | 24-48 hours Post administration of anti-COVID-19 medication | **At the time of discharge | Three-month follow-up | Six-month follow-up |
| 1 | Age (years) | x |  |  |  |  |  |
| 2 | Gender | x |  |  |  |  |  |
| 3 | Weight (kg) | x |  |  |  |  |  |
| 4 | Height (cm) | x |  |  |  |  |  |
| 5 | Smoking status | x |  |  |  |  |  |
| 6 | Date of onset of illness | x |  |  |  |  |  |
| 7 | Fever at the onset of illness | x |  |  |  |  |  |
| 8 | Difficulty in breathing at the onset of illness | x |  |  |  |  |  |
| 9 | Duration of illness (days) | x |  |  |  |  |  |
| 10 | Co-morbid | x |  |  |  |  |  |
| 11 | *** CURB-65 score during hospital stay | x |  |  |  |  |  |
| 12 | *** qSOFA score during hospital stay | x |  |  |  |  |  |
| 13 | ICU stay | x |  |  |  |  |  |
| 14 | Length of stay at ICU | x |  |  |  |  |  |
| 15 | Length of stay at hospital | x |  |  |  |  |  |
| 16 | Travel history | x* |  |  |  | x* |  |
| 17 | History of influenza vaccine | x* |  |  |  | x* |  |
| 18 | Treatment with anti-COVID-19 medicines | x |  |  |  |  |  |
| 19 | Day of illness at which RT-PCR negative | x |  |  |  |  |  |
| 20 | Need of mechanical ventilation | x |  |  |  |  |  |
| 21 | Laboratory investigations | x | x | x | x | x | x |
| 22 | Radiology (CXR and/or CT-Chest) | x | x | x | x | x | x |
| 23 | DLCO/Spirometry |  |  |  |  | x | x |
| 24 | ECHO |  |  |  | x | x | x |

\* if not available in hospital records
\*\* Laboratory and radiology tests will be considered as baseline for further reference in the follow-ups
\*\*\* CURB-65: for assessing pneumonia severity, it ranges from 0 to 5. CURB-score $\geq 3$ labeled as severe pneumonia. qSOFA score: Quick Sequential Organ Failure Assessment score, it ranges from 0 to 3 points. The presence of 2 or more qSOFA points near the onset of infection is associated with a greater risk of death or prolonged intensive care unit stay.

Medical record files of the patients will be reviewed to extract the data on diagnosis and management throughout the hospital stay. Demographic data, presenting complaints, co-morbidities, smoking and travel history will be recorded one time. Moreover, quick Sequential Organ Failure Assessment (qSOFA) and (CURB-65) scores will also be calculated anytime during hospital stay. Conversely, the radiology and laboratory data of 24-72 hours before and after the administration of anti-COVID-19 disease medications (remdesivir, hydroxychloroquine, convalescent Plasma therapy etc.) and steroids will be mentioned. Any missing information from medical record file will be collected directly from patient at the time of follow-up.

### Planning and Training

Necessary approvals from ethical review committees will be taken. Training will be conducted for study staff at the study site (AKUH) by the study investigators. The training will focus on introduction of the project and the team members, administration and completeness of data collection tools including informed consent, communication skills and other study activities. The staff will also be trained on his/her own safety during this outbreak and training on usage of personal protective equipment will also be provided.

### Data Management, Storage and Security

All data will be collected at the study site using the data collection tool specifically developed for the study. Each participant will be assigned unique study identification. Data will be collected on paper-based forms, secured in a locked cabinet at the study site. Data will be entered in SPSS and stored in password protected-user specific computer database. Only the study investigators will have access to the data. If the data entry error rate is >10%, the data will be checked. In case when the error rate is <10%, discrepant entries will be verified and corrected. Daily Log sheet will be maintained by the study staff for screening and enrolling the participants and to maintain their follow-ups. The study staff will maintain a log sheet for performing and recording the lung function test, CXR, ECHO and CT-chest findings.

### Ethical Consideration

The project was approved by the Ethical Review Committee (ERC) of the AKUH (ERC reference number 2020-4735-11311).

### Consent and Assent

Informed consent will be taken from patients, parents, or legal guardians of the child. Purpose of the study as well as all study procedures will be explained in detail to the caregiver/participant. Assent will also be obtained from the children of age 15 years-18 years in addition to consent from their legal caregiver/parents. Verbal consent will be obtained on phone at least 2 weeks after hospital discharge if fulfills the eligibility criteria. Written informed consent/assent will be obtained at three-month visit to a hospital for testing. The process of obtaining consent may take up to 15-20 minutes.

### Risks and Benefits

There are no risks in participating in this study. In rare cases, patient may experience light-headedness/syncope during the pulmonary function test. Patients participating in this study will have access to their investigations to track the progress of their lung functions. Patients will be followed up in the clinic and examined by an ID physician and/or pulmonologist free of cost. All the tests which are part of the study protocol will be performed from the study budget. The study may cover the travel to and from the hospital for participants needing support. No other direct benefits to the individual participant are anticipated.

### Plan of Statistical Analysis

The data will be analyzed using standard statistical software. The main outcome variable is Lung function at 3 and 6 months. Frequencies with percentages will be reported for categorical variables like lung functions, gender, fever at the onset of illness, history of influenza vaccine, category of illness, co-morbid, ICU stay, travel history, smoking status, mechanical ventilation etc. For quantitative variables such as age (years), weight (kg), height (cm), length of hospital stay (days), length of ICU stay (days), duration of mechanical ventilation (days), FEV1, FVC, FEV1/FV ratio, qSOFA score, CURB-65 score, mean/median ± standard deviation/IQR will be reported depending upon the data distribution. Chi-square/fischer exact test will be applied for assessing the association between two categorical variables and student-t-test/ Mann-Whitney U test for the two quantitative variables.

### Study progress

Recruitment to the Lung function COVID-19 study began in August 2020 and is currently ongoing. The study is expected to conclude in March 2021. The current protocol is version 2.0 and is dated 7 July 2020.

## Discussion

There are several strengths and limitations of the study. Recruitment will be using a prospective approach which will be helpful in monitoring the course of the disease in real time and plan and prescribe any intervention as needed. The variables in the study have been selected after thorough literature search and opinion of the experts in the field of infectious diseases. The study team includes a mix of researchers, physicians trained in the field of infectious diseases, coordinators and field staff trained in managing mega projects by maintaining quality standards of data management.

The findings will play crucial role in the development of programs for survivors at population level. The study findings will be disseminated to stakeholders including; community, government and academia. Moreover, strategies to avert consequences of disease such as a rehabilitation programs could be devised based on the study findings.

This is a single center study with a small sample size. The current sample size may not be the representative of population at large however; the study has strong potential to develop evidence on the current state of lung function among survivors. Follow-up at six-months could be a challenge with the unpredictable pandemic situation.

## Data Availability

The datasets generated during the current study will be available from the corresponding author on reasonable request

## Abbreviations

AKUH: Aga Khan University Hospital
RT-PCR: Reverse Transcriptase Polymerase Chain Reaction
DLCO: Diffusion Lung capacity for Carbon Monoxide
ID: Infectious Disease
ECHO: Echocardiogram
ERC: Ethical Review Committee
SARS-CoV-2: Severe Acute Respiratory Syndrome Coronavirus 2
WHO: World Health Organization
MERS-CoV: Middle East Respiratory Syndrome Coronavirus
SOPs: Standard Operating Procedures
ARDS: Acute Respiratory Distress Syndrome
CT: Computed Tomography
COPD: Chronic Obstructive Pulmonary Disease
ATS: American Thoracic Society
qSOFA: quick Sequential Organ Failure Assessment
ICU: Intensive Care Unit
HDU: High Dependency Unit

## Ethics approval and consent to participate

The study has been reviewed and approved by Ethical Review Committee (ERC), Aga Khan University Hospital (ERC reference number 2020-4735-11311). Verbal and written informed consent (from patients/caregivers) will be obtained by study staff at the time of enrollment in the study.

## Consent for publication

Not applicable

## Patient and public involvement

Patients were not involved in the design and conception of this study. Refer to the ‘Methods and design’ section for further details. However, the radiology and laboratory results will be shared with patients at the time of consultation with physician. The overall findings of the study will be disseminated at larger level through presentations in seminars and conferences.

## Author contributions

All listed authors adhere to the authorship guidelines of this manuscript. All authors have reviewed and agreed to publication. SQ and FN conceptualized the study design and methodology, NN and NR provided input as an expert in infectious diseases and pulmonology. FN, SQ, NA and ZH contributed in the write up of manuscript.

## Acknowledgements

The lung function COVID-19 study team would like to thank study participants who have participated in this research to date. Team would also like to acknowledge the efforts of research staff for showing their dedication towards the completion of this project.

